# Excess mortality across regions of Europe during the first wave of the COVID-19 pandemic – impact of the winter holiday travelling and government responses

**DOI:** 10.1101/2020.11.24.20237644

**Authors:** Jonas Björk, Kristoffer Mattisson, Anders Ahlbom

## Abstract

**Background:** This aggregated population study of 219 regions in 11 European countries investigated the effect of the seemingly quasi-randomly assigned school winter holiday week on excess mortality associated with the COVID-19 pandemic during spring 2020. A secondary aim was to evaluate the impact of stringency and timing of the government responses to the early inflow of infected cases.

**Methods:** Regional data on mortality week 14-23 in 2020 compared with the same period 2015-2019 were retrieved from Eurostat and national statistical agencies. Data on initial government responses were obtained from the Oxford COVID-19 Government Response Tracker. Variance-weighted least square regression was used with further adjustment for population density and age distribution.

**Results:** Being a region with winter holiday exclusively in week 9 was in the adjusted analysis associated with 16 weekly excess deaths (95% confidence interval 13 to 20) per million inhabitants, which corresponds to 38% of the excess mortality during the study period in these regions. A more stringent response implemented in week 11, corresponding to 10 additional units on the 0-100 ordinal scale, was associated with 20 fewer weekly deaths (95% confidence interval 18 to 22) per million inhabitants.

**Conclusions:** Travelling during winter holiday in week 9 was an amplifying event that contributed importantly to the excess mortality observed in the study area during the spring 2020. Timely government responses to the resulting early inflow of cases was associated with lower excess mortality.

## Introduction

Early cases of COVID-19 known to be infected in Europe (21 cases by 21 February, week 8, 2020) were linked to two different clusters, one in Bavaria, Germany, and one in Haute-Savoie, France.^1^ On 22 February the Italian authorities reported clusters of cases in Lombardy and additional cases from three other regions in northern Italy.^2^ These initial clusters were all located in close proximity to the Alps, the most extensive mountain range in Europe. Late detection of the index cases generally delayed isolation of further local cases in the region.^1^ Additionally, the large number of tourists at ski resorts in the Alps during the school winter holidays in February and March most likely contributed importantly to the further spread of the virus across Europe. This assertion is supported by genetic analyses of SARS-CoV-2 in for example Denmark and Iceland, which have associated specific haplotypes with travel to ski resort areas in Austria (haplotype A2a2a) and Italy (A2a1).^3, 4^ Travelling during public holidays has further been found to be important determinants of the COVID-19 transmissions out from the virus epicentres in Wuhan, China, and Qom, Iran.^5-8^

In the present study we focus on 11 countries in Europe that all have school winter holidays and were not part of the initial epicentre: Germany, France, UK, Belgium, Netherlands, Luxemburg, Sweden, Denmark, Norway, Finland and Iceland. Striking regional differences in excess mortality during March – June 2020 have been observed for these countries (Figure 1A).^9^ It is not known, however, to what extent the exact week of the school winter holiday, which varied from week 6 (early February) to week 10 (early March) across regions,^10^ (Figure 1B), has contributed to the variation in mortality. We hypothesized that the exact week of the winter holiday was not only important for the initial spread of the disease, but also essentially quasi-random with respect to structural determinants of the further development of the pandemic and its effect on the mortality across regions.

**Figure 1A.**
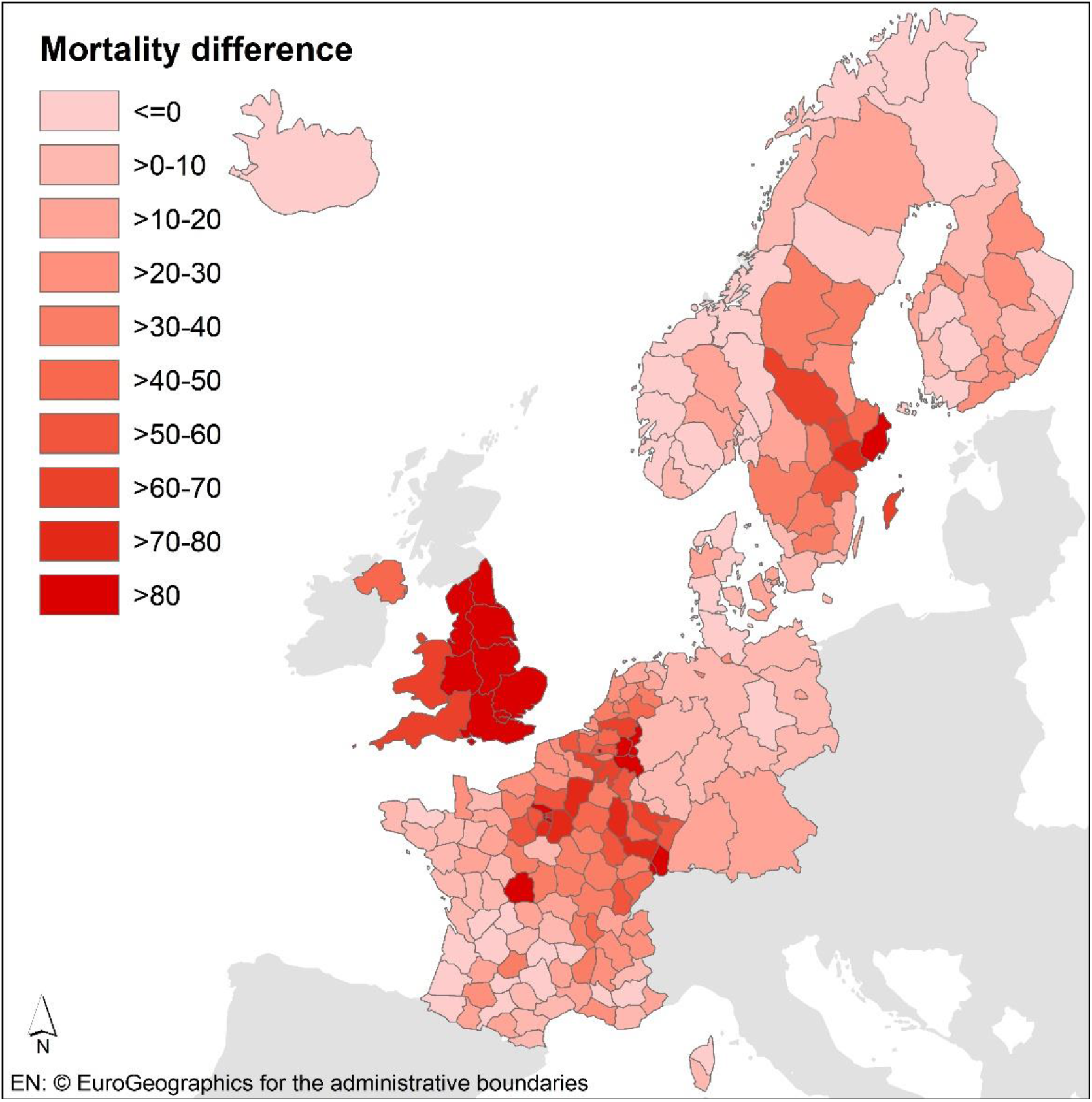
Differences in weekly all-cause mortality per 1000 000 inhabitants in 219 European regions during week 14 to 23 in 2020 and the same period 2015 – 2019 (2016 – 2019 for Germany and Netherlands)

**Figure 1B.**
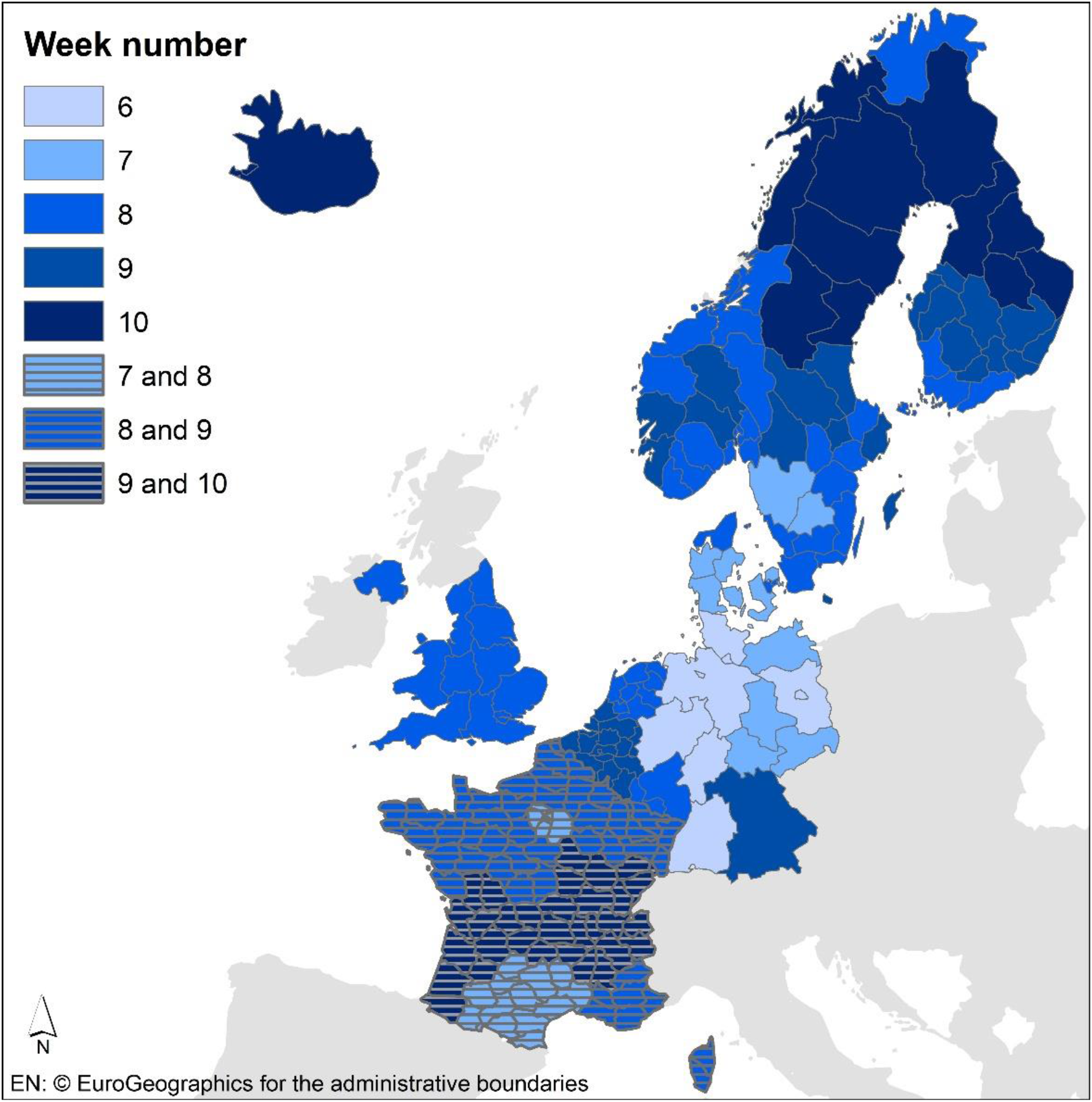
School winter holiday week (6 to 10) in the 219 regions countries included in the study.

The governments in these 11 European countries reacted differently and with different timing in the implementation of non-pharmaceutical interventions at the population level as a response to the COVID-19 pandemic during the spring 2020.^11^ As example of the differences in response, France and Germany, reacted stringently and early, UK reacted stringent but with a delay, while Sweden maintained a less stringent reaction throughout this period (Figure 2). Previous studies have suggested that a timely response in relation to when initial infections occurred had critical and long-term effects.^12, 13^ However, so far no systematic effort has been made to quantify this effect on mortality while taking the influence of amplifying events or other population-level factors into account.

**Figure 2.**
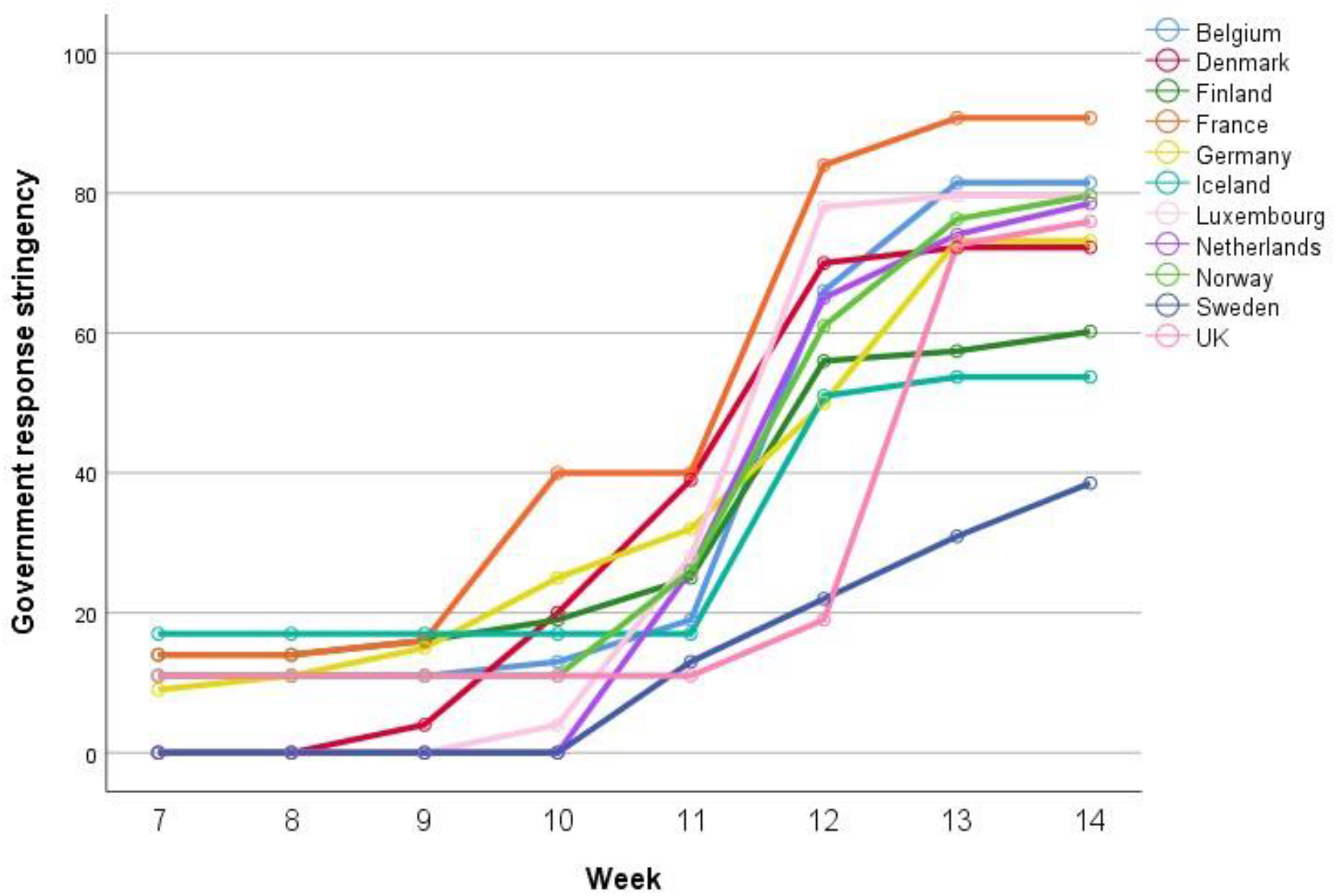
Stringency in the government response (index 0 – 100) during week 7 – 14 in the 11 countries included in the study

The overall aim of the present aggregated population study of 219 regions in 11 European countries was to exploit the effect of the seemingly quasi-randomly assigned school winter holiday week on the excess mortality during spring 2020. As a secondary aim, the impact of stringency and timing of the initial government responses was evaluated.

## Methods

### Study population

The present study includes population data 2015 – 2020 for 219 regions in 11 European countries (B, D, DK, F, FIN, ISL, LUX, N, NL, S and UK) that generally all have school winter holidays for at least one week. An underlying assumption when defining the study area was that a significant part of the population having winter holiday in the included countries travel to the Alps for skiing,^14^ including the areas in Austria and northern Italy affected early during the COVID-19 pandemic.

### Data collection

Regional data for the present study were originally extracted from European Data Journalism Network,^9^ and subsequently verified and updated by data from Eurostat and national statistical agencies. Most data were gathered on the NUTS (Nomenclature des Unités Territoriales Statistiques) level 3 (169 regions), but Germany and UK only had mortality data published on NUTS 1 (16 and 11 included regions, respectively) and Netherlands only on NUTS 2 (12 regions). We therefore extracted data for the neighbouring country Belgium on the same level (NUTS 2; 11 regions) for comparability reasons. Number of deaths during week 14 (30 Mar – 05 Apr 2020) until week 23 (01-07 Jun 2020) were retrieved for the different regions together with the annual average number of deaths during the same period 2015 – 2019 (2016 – 2019 only available for Germany and the Netherlands). We projected population size of each region linearly for 1 January 2020 based on size 2015 – 2019. Average population size 2015 – 2019 was calculated from data on size 1 January each year.

Data on school winter holiday weeks for each study region were directly accessible from a Swiss website ^10^, and verified for a subsample. Nine regions in Germany either did not have a full week winter holiday (n = 7), or had the holiday already week 6 (n = 2). All other 210 regions had their winter holiday between week 7 and 10 (Table 1 and Figure 1B). France had two-week winter holidays (week 7 + 8, 8 + 9 or 9 + 10). The proportion of the study population exposed to winter holidays the different weeks in each country is presented in Supplementary Table S1. Scotland (UK NUTS 1) was not included in the study as its winter holiday week varied on finer geographical levels than available mortality data. Rest of UK had their winter holiday solely in week 8 and Belgium solely in week 9.

**Table 1.** Basic characteristics of the 219 European regions included in the study, stratified by the timing of the school winter holiday.

|  | All | No winter holiday<br>or week 6 | Week 7,<br>7 to 8<sup>b</sup> | Week 8,<br>8 to 9<sup>b</sup> | Week 9,<br>9 to 10<sup>b</sup> | Week 10 |
| --- | --- | --- | --- | --- | --- | --- |
| Number of<br>regions | 219 | 9 | 13<br>21 | 51<br>43 | 37<br>32 | 13 |
| Countries | B, D, DK, F, FIN,<br>ISL, LUX, N, NL,<br>S, UK | D | D, DK, S; F | D, DK, FIN,<br>LUX, N, NL, S,<br>UK; F | B, D, DK, FIN, N,<br>NL, S; F | FIN, N, ISL, S |
| Population size,<br>median (range,<br>million) |  | 3.7 (0.69 – 18) | 0.80 (0.37 – 4.1)<br>0.75 (0.08 – 2.2) | 0.55 (0.03 – 9.2)<br>0.60 (0.14 – 2.6) | 0.38 (0.04 – 13)<br>0.41 (0.12 – 1.9) | 0.23 (0.07 – 0.41) |
| Population<br>density, median<br>(range) | 86 (1 – 21 040) | 312 (86 – 4 290) | 109 (34 – 4 480)<br>129 (15 – 21 040) | 134 (2 – 5 690)<br>99 (24 – 453) | 54 (87.9 – 7 470)<br>62 (21 – 576) | 7 (1 – 218) |
| *Total* |  |  |  |  |  |  |
| Inhabitants,<br>(million) | 267 | 55.3 | 16.6<br>18.2 | 88.0<br>29.9 | 39.6<br>16.8 | 2.7 |
| Proportion below<br>20 years, % | 21.8 | 18.7 | 19.1<br>24.7 | 22.9<br>23.8 | 21.0<br>23.1 | 22.5 |
| Proportion above<br>70 years, % | 14.4 | 15.4 | 16.8<br>12.5 | 13.4<br>14.9 | 14.2<br>15.7 | 14.9 |
| Response index<sup>a</sup><br>week 11, median<br>(range) | 32 (11 – 40) | 32 (32 – 32) | 39 (13 – 39)<br>40 (40 – 40) | 26 (11 – 39)<br>40 (40 – 40) | 25 (13 – 39)<br>40 (40 – 40) | 25 (13 – 26) |

<sup>a</sup>Stringency in the non-pharmaceutical interventions on the population level (index 0 to 100) obtained from the Oxford COVID-19 Government Response Tracker <sup>11</sup>. All regions within a country were assigned the same value.
<sup>b</sup>Second row represents French regions that all have two weeks of winter holiday

Daily country-level data on initial non-pharmaceutical interventions at the population level were obtained from the Oxford COVID-19 Government Response Tracker.^11^ Specifically, we used their stringency index (0 – 100) which is an average of 9 ordinal items, school closing, work place closing, cancelation of public events, restrictions on gatherings, closing of public transports, stay at home requirements, restrictions on internal movement, international travel controls and public information campaigns. We calculated weekly averages for each country during week 8 (17 – 23 Feb) to week 14 (30 Mar – 5 Apr) based on data for Monday through Friday (Figure 2). No specific regional data on government response were available, which implies that all regions within a country were assigned the same value for the stringency index a given week. We considered this appropriate given that the initial responses were mainly acting on national rather than on regional levels.

### Statistical analysis

Statistical analyses were conducted in Stata SE 14.2 (Stata Corp.) and IBM SPSS Statistics 25 (SPSS Corp.). We calculated the mortality difference (MD) for week 14 to 23 for each region

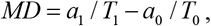

where *a*_*1*_ represents the number of deaths that occurred in this period during 2020, *a*_*0*_ the average number of deaths in the same period during 2015-2019 (2016-2019 for Germany and Netherlands), *T*_*1*_ represents the number of person-weeks of follow up in 2020, and *T*_*0*_ the average number of person-weeks in the comparison years. The number of person-weeks each year was calculated as the population size at 1 January multiplied by 10 weeks of follow up. Thus, MD represents the excess death rate, and was reported as the number of extra deaths per million inhabitants and week.

MD was first presented for each region stratified by the winter holiday week. The association between winter holiday week and the mortality difference (2020 vs. 2015 – 2019) in each region was investigated further using linear regression with variance-weighted least squares (*vwls* command in Stata). The observed MD for each region was in all regression analyses weighted with the inverse of its estimated variance

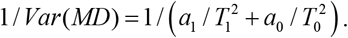

The weighted analysis implies that observations for regions with larger population sizes, and thus more precise estimates of MD, are more influential than those for smaller regions with fewer inhabitants. Holiday week was used as categorical in the analysis with regions with no winter holiday or holiday in week 6 (all in Germany) as a collapsed reference category due to small numbers. The two-week holidays in France were classified by placing one half of the deaths and person-weeks in the first week and the other half in the second week. The regression analysis for the association between winter holiday week and MD was presented both unadjusted and with adjustment for country (categorical without reference category), the population density of the region on the log-scale, the proportions of individuals below 20 (most of them exposed to school holidays) and the proportion above 70 years of age (as an indicator of the vulnerability of the population).

In separate linear regression analyses, we investigated the association between country-level stringency of the government response (index 0 – 100) during week 8 – 14, one week at a time, and the regional MD for the entire follow up. The dependent variable, holiday-, density and age-adjusted MD, and its variance-weights were obtained from the previous analytical step. We also estimated the percentage of the observed country-level differences in mortality that could be attributed to regional differences in winter holiday week, population density and government response stringency, respectively.

## Results

A total of 586 977 deaths occurred in the study area during week 14 – 23 in 2020, which is 105 230 (+22%) more deaths than expected from the average death rate during the same period in 2015 – 2019. This corresponds to 39 excess deaths (MD 39.2, 95% confidence interval [CI] 38.4 to 39.9) per million inhabitants and week.

Elevated MDs were observed in non-French regions having their winter holidays in week 8 or 9, and for the French regions irrespectively of the timing of winter holiday (Table 2 and Figure 3). There were differences in population density, age distribution and country affiliation between regions that had their winter holiday early (week 6 – 7), in the middle (week 8 – 9) or late in the season (week 10; Table 1). In particular, regions having winter holiday week 9 or 10 were generally less dense than regions having holiday earlier or not at all. Thus, adjustments for population factors led to marked changes in the estimate effects associated with winter holiday week (Table 3, Model 1 and 2). In the regions outside France, having winter holiday in week 9 was in the adjusted analysis associated with 16 excess deaths (MD 16.3, 95% CI 12.7 to 20.0; Table 3, Model 3) per million inhabitants and week compared to the reference (winter holiday in week 6 or no holiday at all). By contrast, no apparent effect of winter holiday week on mortality was observed for the French regions (Table 3, Model 4). Population density of the region was clearly associated with mortality. A tenfold higher population density (e.g. 1 000 rather than 100 inhabitants per km^2^) was associated with 12.5 and 20.2 excess deaths per million habitants and week in the non-French and French regions, respectively. Age distribution of the population was essentially unrelated to the regional excess mortality.

**Table 2.** All-cause weekly mortality per million inhabitants in 219 European regions in 2020 week 11 – 23 compared with the same period 2015 – 2019 (2016 – 2019 for Germany and Netherlands), grouped by the timing of the school winter holiday.

|  | 2020 |  |  | 2015-2019 |  |  |  |
| --- | --- | --- | --- | --- | --- | --- | --- |
| Winter holiday, week | Deaths, n | Person-weeks, million | Death rate | Deaths, n | Person-weeks, million | Death rate | MD<sup>a</sup> (95% CI) |
| No winter holiday or week 6 | 117 499 | 552.8 | 213 | 110 969 | 543.1 | 204 | 8.2 (6.5 to 9.9) |
| Week 7 | 37 996 | 165.6 | 229 | 36 388 | 164.2 | 222 | 7.9 (4.6 to 11.1) |
| Week 7 to 8<sup>b</sup> | 34 546 | 182.3 | 190 | 24 532 | 180.1 | 136 | 53.3 (50.7 to 55.9) |
| Week 8 | 210 432 | 880.5 | 239 | 146 432 | 862.0 | 170 | 69.1 (67.8 to 70.5) |
| Week 8 to 9<sup>b</sup> | 61 319 | 299.2 | 205 | 53 199 | 298.8 | 178 | 26.9 (24.7 to 29.1) |
| Week 9 | 86 749 | 396.4 | 219 | 68 744 | 389.2 | 177 | 42.3 (40.3 to 44.2) |
| Week 9 to 10<sup>b</sup> | 33 196 | 168.4 | 197 | 29 679 | 167.1 | 178 | 19.5 (16.6 to 22.5) |
| Week 10 | 5 240 | 27.5 | 191 | 4 995 | 27.2 | 184 | 6.8 (-0.4 to 14.1) |
| Total | 586 977 | 2 6762.6 | 220 | 474 926 | 2631.7 | 180 | 39.2 (38.4 to 39.9) |

<sup>a</sup>Mortality difference, i.e. difference between death rate in 2020 and 2015-2019
<sup>b</sup>French regions have two weeks of winter holiday

**Table 3.** Weighted multivariable linear regression analysis for the association between school winter holiday week and the all-cause mortality difference (MD) per week and million inhabitants in 219 European regions, contrasting 2020 week 14 – 23 with the same period in 2015 – 2019 (2016 – 2019 for Germany and Netherlands). Results are presented unadjusted and adjusted for population density, age distribution (proportion of individuals below 20 and above 70 years of age) and country.

|  | All regions (n = 219) |  | Non-French regions (n = 123) | French regions (n = 86) |
| --- | --- | --- | --- | --- |
| Variable | Model 1.<br>Unadjusted results<br>MD (95% CI) | Model 2.<br>Adjusted results<br>MD (95% CI) | Model 3.<br>Adjusted results<br>MD (95% CI) | Model 4.<br>Adjusted results<br>MD (95% CI) |
| Constant | 8.6 (6.9 to 10.3) | - | - | 23.9 (17.2 to 30.6) |
| Winter holiday |  |  |  |  |
| No, or week 6 | Ref. | Ref. | Ref. | - |
| Week 7 | 20.0 (17.1 to 23.0) | 13.4 (9.7 to 17.0) | 2.6 (-2.0 to 7.1) | Ref. |
| Week 8 | 51.0 (48.9 to 53.0) | 5.9 (2.3 to 9.5) | -2.9 (-7.1 to 1.2) | -3.2 (-7.7 to 1.4) |
| Week 9 | 26.1 (23.8 to 28.4) | 13.1 (9.7 to 16.5) | 16.3 (12.7 to 20.0) | -6.2 (-11.2 to -1.3) |
| Week 10 | 7.8 (3.9 to 11.8) | 6.2 (1.0 to 11.4) | 4.2 (-5.5 to 13.9) | -7.5 (-13.5 to -1.5) |
| Log10(Density) | - | 17.3 (14.9 to 19.8) | 12.5 (9.2 to 15.8) | 20.2 (16.4 to 23.9) |
| Proportion below 20 | - | 2.3 (1.6 to 3.1) | 1.1 (-0.11 to 2.2) | 0.92 (-0.27 to 2.1) |
| Proportion above 70 | - | -0.32 (-1.0 to 0.40) | 0.17 (-0.74 to 1.1) | -2.1 (-3.3 to -0.83) |

Log10(Density) centered at 100 persons per km<sup>2</sup>, proportion below 20 and above 70 at 20% and 15%, respectively

**Figure 3.**
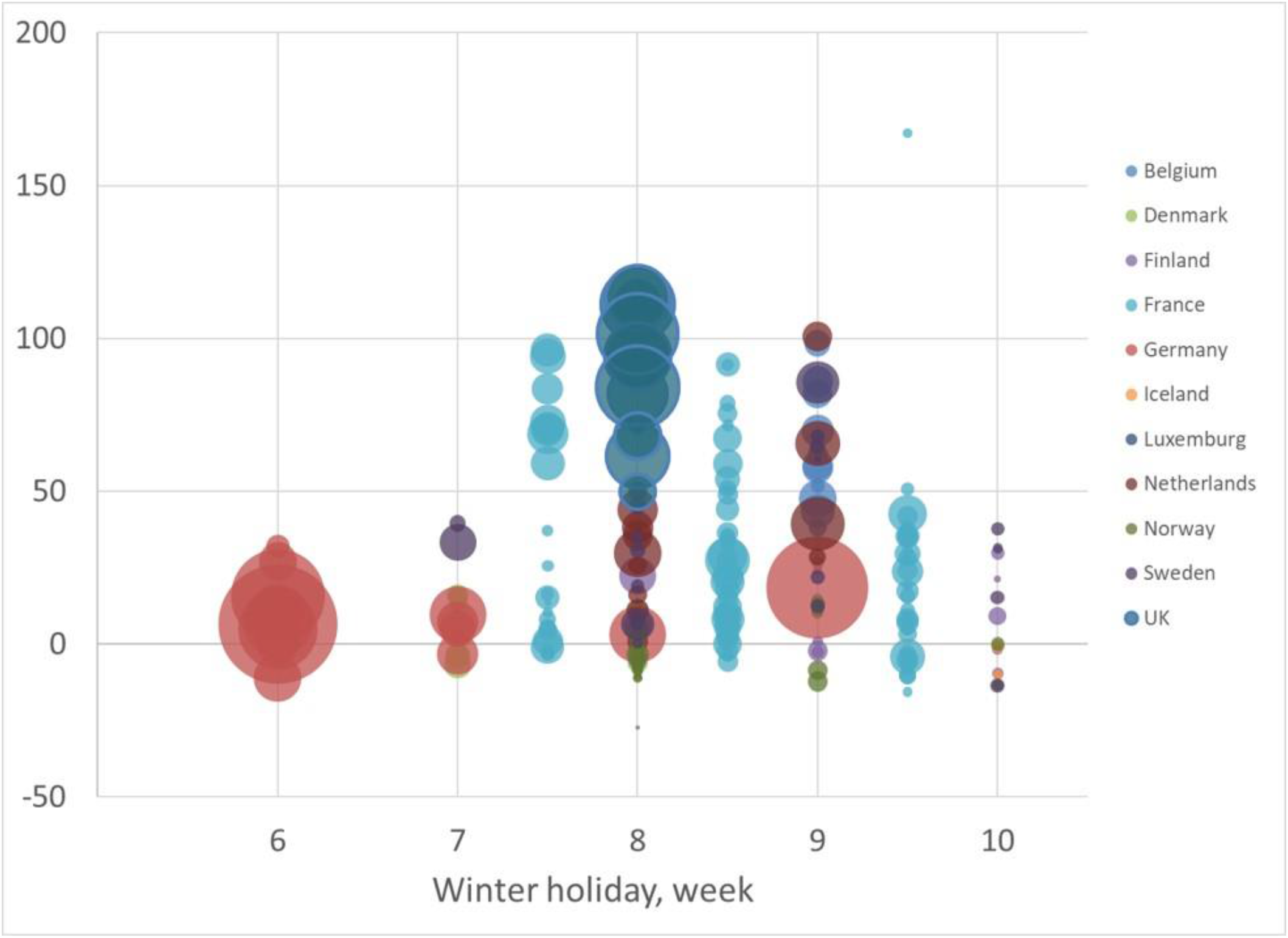
All-cause mortality difference (MD) per week and million inhabitants in 219 European regions, contrasting 2020 week 14 – 23 with the same period in 2015 – 2019 (2016 – 2019 for Germany and Netherlands), and stratified by the timing of the school winter holiday (week 6 to 10). Regions with no winter holiday were collapsed with week 6. The French regions with two weeks winter holiday were placed in between the first and second week. The area of the bubble is proportional to population size.

Overall, an estimated 6.1% (95% CI 4.8 to 7.5%) of the excess deaths in the study area was attributed to having winter holiday in week 9, but this proportion varied substantially across different countries. Large number of excess deaths was attributed to winter holiday week 9 in the regions of Germany (Bavaria), Belgium (all 11 regions), Netherlands (4 regions) and Sweden (Stockholm and 5 other regions; Table 4). By contrast, the winter holiday week did not seem to explain any of the excess mortality in UK or France. As a comparison, 19% of the excess mortality (95% CI 14 to 24) was attributed to population density above 100 persons per km^2^. Population density explained large proportions of the excess mortality in Germany (66%) and France (27%) but only a minor proportion in for example Sweden (4%; Table 4).

**Table 4.** Proportion of the all-cause mortality difference (MD) per week and million inhabitants in 11 European countries, contrasting 2020 week 14 – 23 with the same period in 2015 – 2019 (2016 – 2019 for Germany and Netherlands), that was attributed to school winter holiday in week 9, population density above 100 persons per km^2^ and a response stringency index below 25 in week 11

| Country | MD (95% CI) | Attributable proportion, % (95% CI) |  |  |
| --- | --- | --- | --- | --- |
|  |  | Winter holiday week 9 | Density above 100 per km<sup>2</sup> | Response<sup>b</sup> below average week 11 |
| Belgium | 63.2 (59.5 to 66.9) | 26 (20 to 32) | 15 (11 to 19) | 18 (16 to 20) |
| Denmark<sup>a</sup> | 1.7 (-3.1 to 6.4) | - | - | - |
| Finland | 12.3 (7.3 to 17.3) | 52 (40 to 64) | 8.6 (6.3 to 11.3) | 0 |
| France | 33.8 (32.4 to 35.3) | 0 | 27 (22 to 32) | 0 |
| Germany | 9.7 (8.3 to 11.1) | 28 (21 to 34) | 66 (48 to 86) | 0 |
| Iceland<sup>a</sup> | -3.7 (-20.3 to 12.8) | - | - | - |
| Luxembourg | 8.7 (-4.1 to 21.5) | 0 | 55 (41 to 73) | 0 |
| Netherlands | 40.6 (37.8 to 43.4) | 17 (13 to 21) | 23 (17 to 31) | 0 |
| Norway<sup>a</sup> | -2.9 (-7.4 to 1.5) | - | - | - |
| Sweden | 42.3 (38.6 to 46.0) | 13 (10 to 16) | 4.1 (3.0 to 5.4) | 55 (49 to 61) |
| United Kingdom | 91.7 (90.1 to 93.4) | 0 | 10 (7.2 to 12.9) | 30 (26 to 33) |
| Total |  | 6.1 (4.8 to 7.5) | 19 (14 to 24) | 20 (17 to 22) |

<sup>a</sup>Attributable proportions were not estimated for countries with no or low (< 5) excess mortality
<sup>b</sup>Stringency in the non-pharmaceutical interventions at the population level (index 0 to 100) obtained from the Oxford COVID-19 Government Response Tracker <sup>11</sup>.

Timing and stringency of the government response in relation to excess mortality was illustrated for all 219 regions and the subset of 38 regions with winter holiday in week 9 in Figure 4. Response data were analysed further on the country-level after adjustment for winter holiday week, population density and age distribution of the regions (Supplementary Table S2). A more stringent response implemented in week 11, corresponding to 10 additional units on the 0 – 100 ordinal scale, was associated with 20 fewer deaths (95% CI 18 to 22) per million inhabitants and week. Weaker associations with excess mortality were in the adjusted analyses observed for responses in week 9 – 10 and 12. The associations with responses implemented early (week 8) or late (week 13 – 14) were only marginal. Overall, 20% of the excess mortality (95% CI 17 to 22%) in the study area was attributed to response stringency in week 11 that was below average (Table 4). For Sweden, we estimated that 55% of the excess mortality (95% CI 49 to 61%) was associated with a less stringent response.

**Figure 4.**
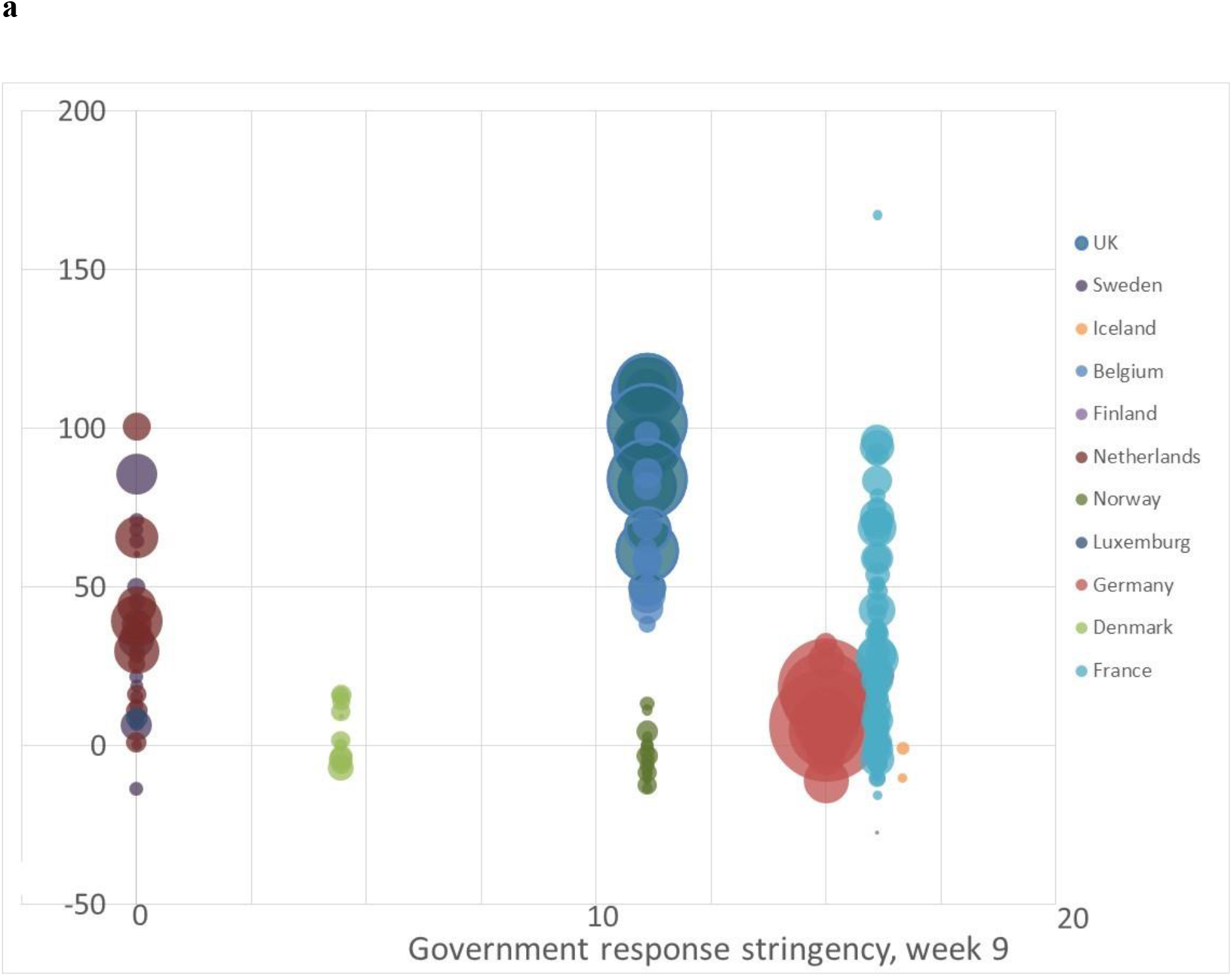
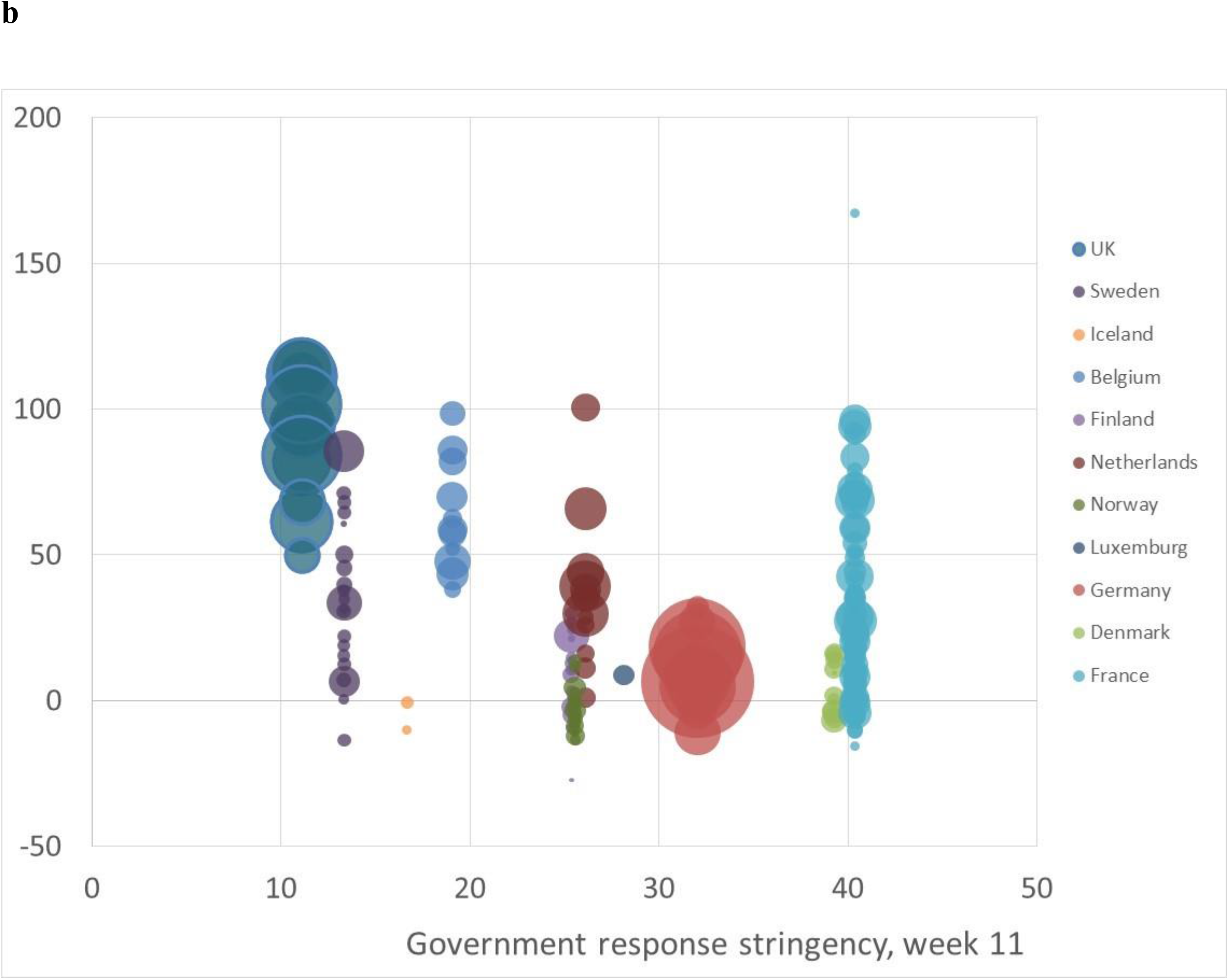
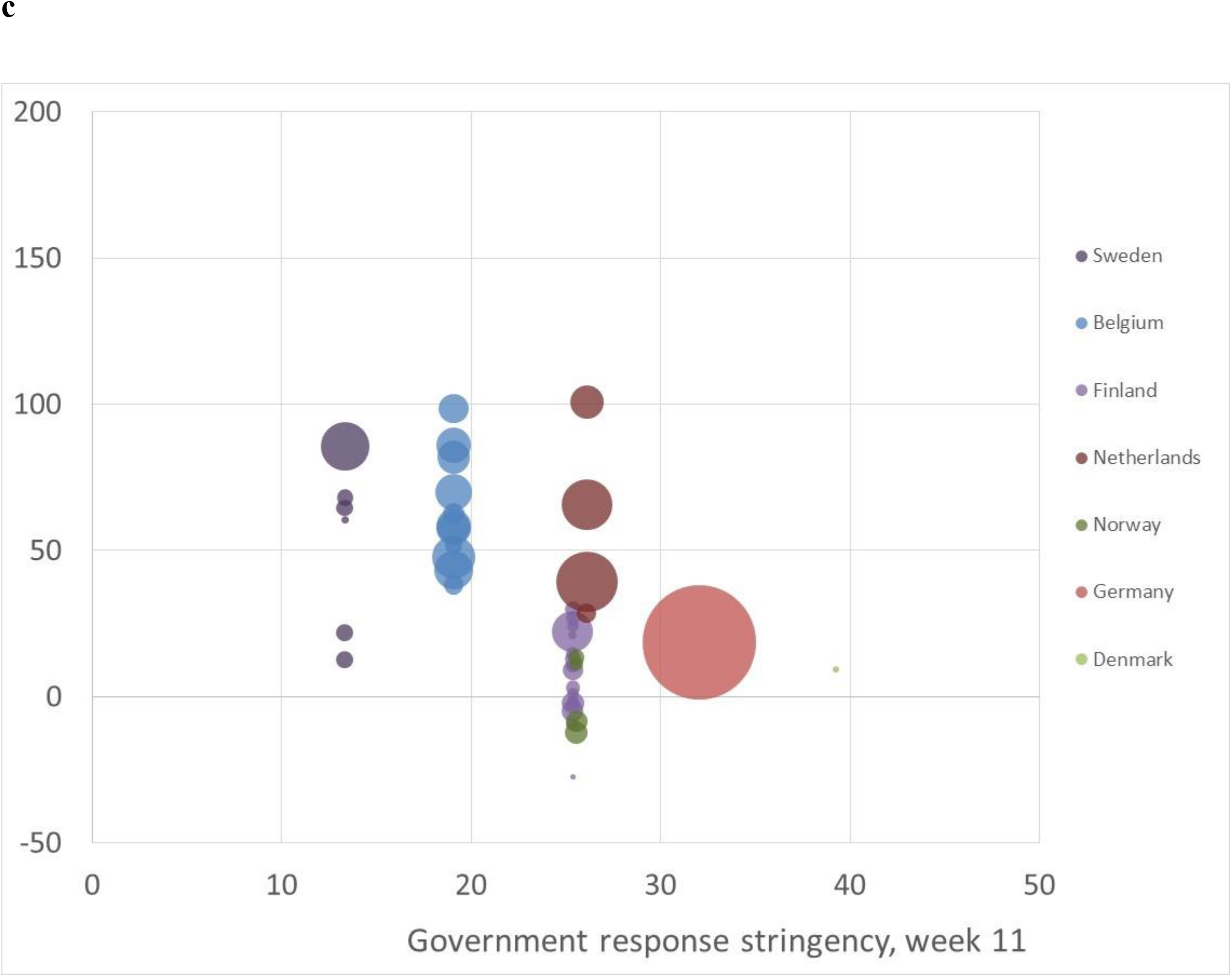
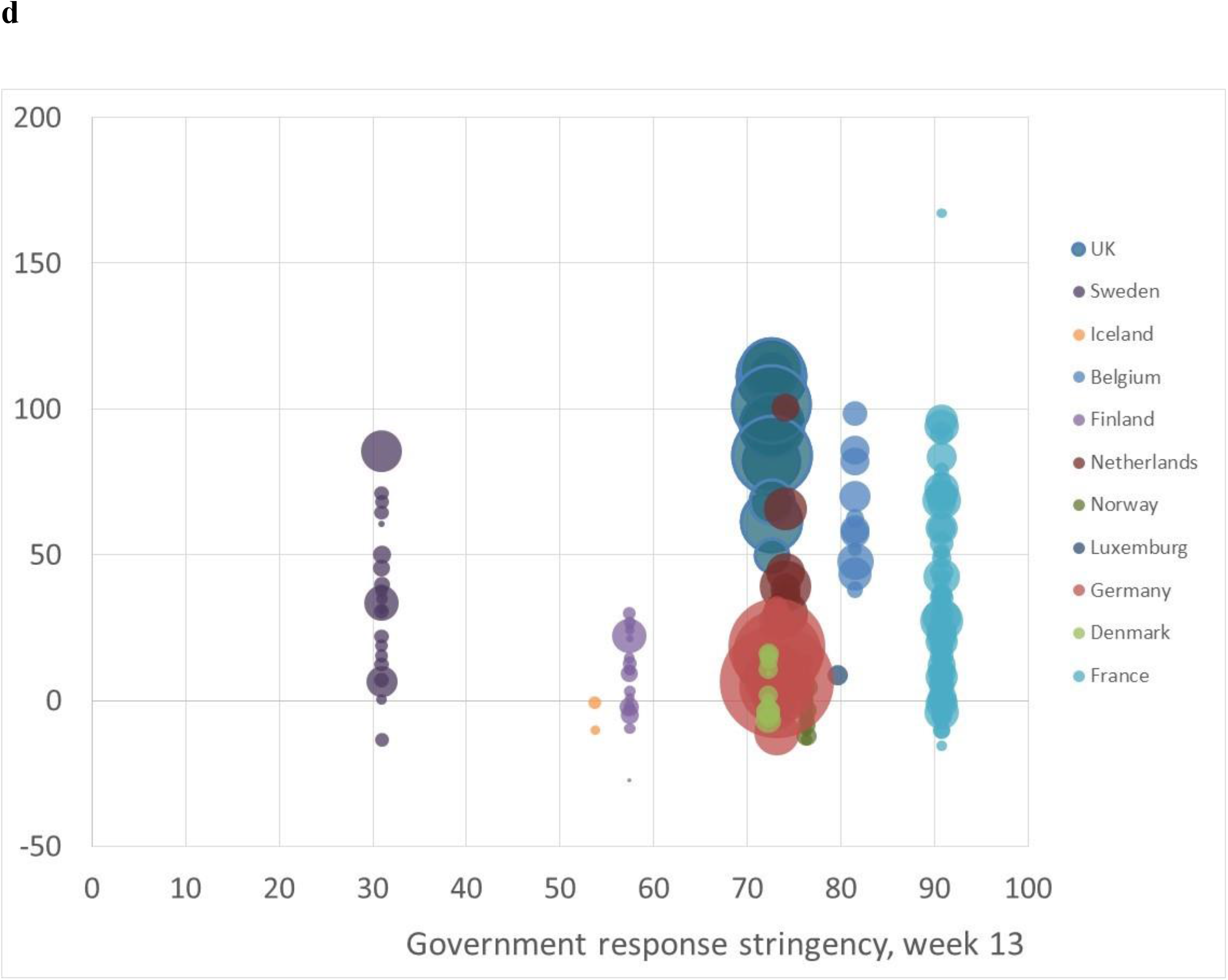
All-cause mortality difference (MD) per week and million inhabitants in 219 European regions, contrasting 2020 week 14 – 23 with the same period in 2015 – 2019 (2016 – 2019 for Germany and Netherlands), and stratified by the government response stringency (index 0 – 100) in each country **a** Week 9, **b** Week 11, **c** Week 11 restricted to the 38 regions with school winter holiday in week 9, **d** Week 13. The area of the bubble is proportional to population size.

## Discussion

A salient finding of the present study was that the timing of winter holiday contributed importantly to the excess mortality observed in the investigated 219 European regions during the spring 2020. The winter holiday was held in week 9 in 38 regions constituting one fifth of the study population, and was in these regions estimated to explain 38% of the excess mortality. Another important finding was that a timely government response to the resulting early inflow of cases was associated with lower excess mortality. By contrast, the government responses early in the pandemic (week 8) or late when the transmission was already widespread (week 13) did not seem to explain any of the observed differences in excess mortality.

No effect of the winter holiday on the mortality was discerned for the French regions despite that the majority (75 out of 96 regions) had school holiday that included week 9. A possible explanation could be that the French population do not go abroad on winter holiday to the same extent as other European countries.^15^ The proportion of trips in Q1 (first quarter) with at least one night stay with an international destination, was only 10% in France, compared to for example 73% in Belgium, 29% in Denmark and 27% in Germany and Sweden according to the most recent European quarterly travel statistics in 2011.^15^. Not only the relative but also the absolute number of persons travelling abroad in relation to population size was low in France compared with the other countries.^15^ UK citizens travel abroad during the winter to a large extent (36% of all trips in 2011 Q1) but had school holiday already in week 8, i.e. one week earlier than the peak inflow of cases returning from the Alps. The number of international visitors to the densely populated metropolitan regions of London and Paris is large also during the winter,^14^ which thus seems as a more likely explanation for the for the initial spread of the pandemic in France and UK than returning travellers from the Alps.

Comparisons between countries and regions are essential in order to improve understanding on why some have markedly higher burdens of COVID-19 mortality than others,^16^ but it is equally important that such comparisons are scientifically sound and backed by empirical data. Non-pharmaceutical interventions represent a broad range of individual, environmental and population-related measures that are often implemented simultaneously in a country or region. As a group they have been found to play a critical role in reducing the direct public health impact of COVID-19 in Europe,^17^ but the evidence for the effect of specific public health measures is generally weaker. As example, a rapid Cochrane review on quarantine alone or in combination with other public health measures concluded that the current evidence regarding effects on incident cases and deaths is limited because most studies on COVID‐19 are based on mathematical modelling studies.^18^ A particular strength of our study was that we were able to estimate the effect of government response while controlling for the regional differences in initial spread caused by the timing of the winter holiday, and for differences in population density and age structure. The results confirmed the importance of a timely response to the COVID-19 pandemic suggested by previous real-world studies.^12, 13, 19^ Belgium, Sweden and UK stood out as the three countries in our study area that, according to our estimates, may have experienced lower excess mortality by intervening earlier. The slow response in Belgium may thus explain an important part of the excess mortality compared with its neighbouring country Netherlands. Similarly, a slow and less stringent response in Sweden may explain an important part of the excess mortality compared with the neighbouring Nordic countries.

Cross-national differences in socioeconomic structures has been put forward as a potential explanation for some of the observed differences in excess mortality during the COVID-19 pandemic.^20^ A population-based study from Sweden with follow up until early May, 2020, found similar socioeconomic gradients in COVID-19 mortality as for mortality from all other causes.^21^ Additionally, an elevated mortality was observed among immigrants from low- and middle-income countries also after adjusting for socioeconomic characteristics. There are differences in this socioeconomic gradient in health across the Nordic countries, but neither the socioeconomic structure nor the strength of the gradient appear to be particularly disadvantageous in Sweden.^22^ Thus, higher population vulnerability is not likely to be an important explanation for the excess mortality observed in Sweden compared to its neighbours.

Our study had several limitations. The use of aggregated data means that results can be subject to ecological inference fallacy, i.e. that the observed associations are not necessarily reflecting true or correctly estimated associations on the individual level.^16^ The ecological analysis is generally sensitive to choice of adjustment factors. We decided to include adjustment for country, which effectively implies that associations between winter holiday week and excess mortality in each country are pooled together in the regression analysis. But it also means that only countries where the winter holiday week differed across regions contribute to the estimated exposure effect. Another limitation was that the calculation of the expected mortality in a region was only adjusted for changes in population size, and did not take time trends, yearly fluctuations (e.g. mean temperature a given year or severity of the flu season) or other population changes into account.^13^ A further limitation was that we that we only estimated the effect of the winter holiday for each region separately and did not consider spill-over effects to neighbouring regions from for example commuting or regional travelling. In this respect, it is therefore likely that our estimates represent the lower bound of the winter holiday effect. It should also be noted that government response was only evaluated in the initial phase of the pandemic. Lockdown measures and other restrictions that last for longer time periods may have substantial adverse health, psychosocial and financial consequences,^17, 23-25^ which were not assessed in our study.

## Conclusion

The timing of the winter holiday and the associated travelling abroad contributed importantly to variation in the excess mortality observed in the study area during the spring 2020. It is important that authorities respond quickly when such amplifying events are identified in order to limit their public health impact.

## Data Availability

The present study was based on secondary, aggregated data that are hosted and maintained by the Lund University Population Research Platform (LUPOP), and can be made available on request. https://www.lupop.lu.se/

## Authors’ contributions

JB conceived the study with a design that was further developed together with AA and KM. JB did the data acquisition and analysis with assistance regarding the mapping from KM. JB drafted the first version of the manuscript, where after all authors revised it critically. All authors have read and approved the final manuscript and agree to be accountable for all aspects of the work.

## Data sharing

The present study was based on secondary, aggregated data that are hosted and maintained by the Lund University Population Research Platform (LUPOP), and can be made available on request. You can find contact information for the data host at https://www.lupop.lu.se/

## Acknowledgments

We want to acknowledge the European Data Journalism Network who collated and published regional data from Eurostat and national statistical agencies that formed the basis for our study.

## Supplementary Tables

**Table S1.**
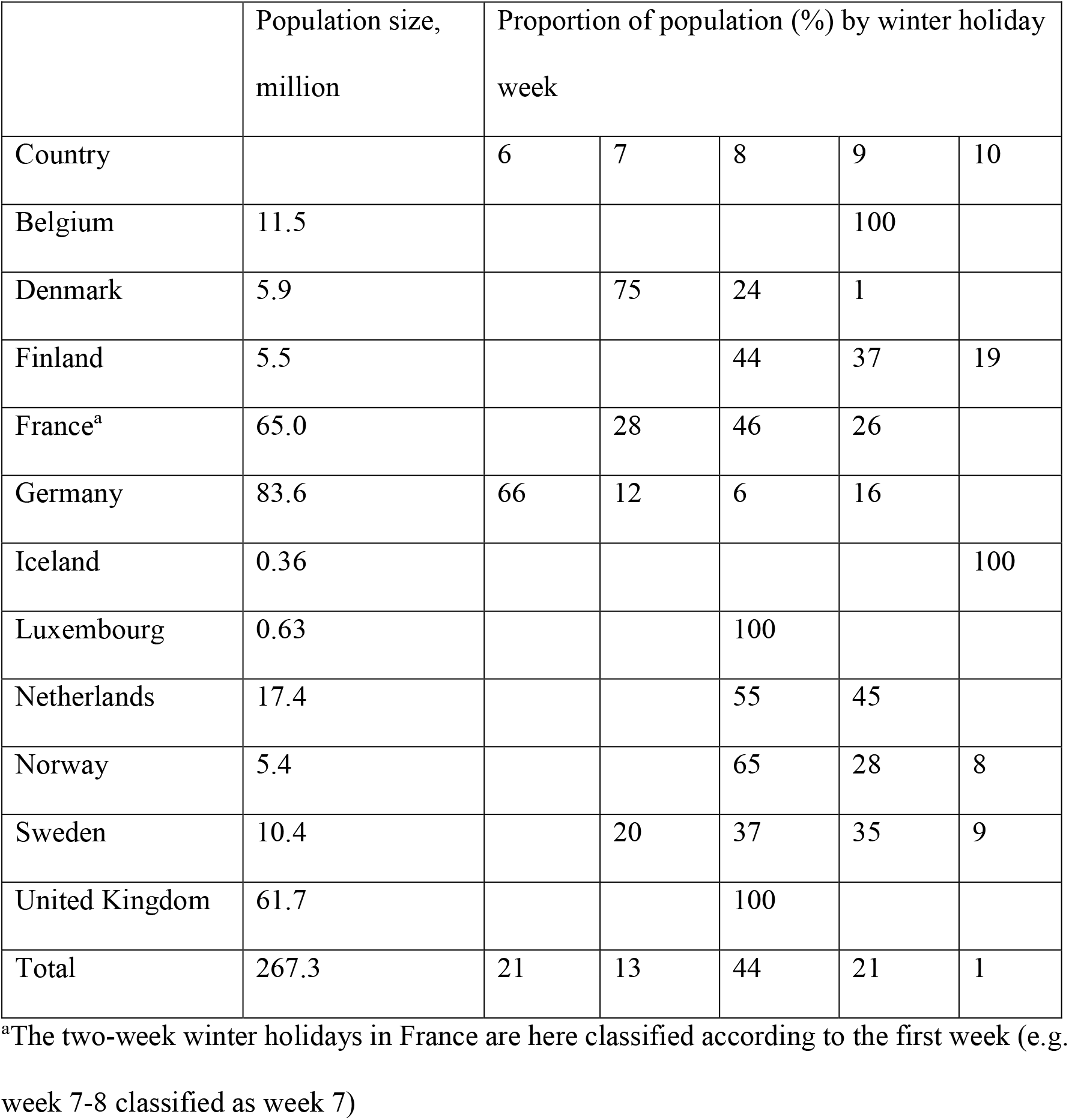
Population size in each country, stratified by the school winter holiday week (6 – 10) of the region they live in.

|  | Population size,<br>million | Proportion of population (%) by winter holiday<br>week |  |  |  |  |
| --- | --- | --- | --- | --- | --- | --- |
| Country |  | 6 | 7 | 8 | 9 | 10 |
| Belgium | 11.5 |  |  |  | 100 |  |
| Denmark | 5.9 |  | 75 | 24 | 1 |  |
| Finland | 5.5 |  |  | 44 | 37 | 19 |
| France<sup>a</sup> | 65.0 |  | 28 | 46 | 26 |  |
| Germany | 83.6 | 66 | 12 | 6 | 16 |  |
| Iceland | 0.36 |  |  |  |  | 100 |
| Luxembourg | 0.63 |  |  | 100 |  |  |
| Netherlands | 17.4 |  |  | 55 | 45 |  |
| Norway | 5.4 |  |  | 65 | 28 | 8 |
| Sweden | 10.4 |  | 20 | 37 | 35 | 9 |
| United Kingdom | 61.7 |  |  | 100 |  |  |
| Total | 267.3 | 21 | 13 | 44 | 21 | 1 |

<sup>a</sup>The two-week winter holidays in France are here classified according to the first week (e.g. week 7-8 classified as week 7)

**Table S2.** Weighted linear regression for the association between government response stringency index (0 – 100) during each week from 8 to 14 and adjusted mortality difference (week 14 to 23, year 2020 vs. 2015 – 2019 ^a^) per week and million inhabitants in 11 European countries

|  |  | Regression coefficients (95% CI) |  |
| --- | --- | --- | --- |
| Week | Response index<sup>b</sup>,<br>mean (range) | Constant, mean<br>centred | Slope per 10 |
| 8 | 8 (0 – 17) | 15 (13 to 17) | -3.4 (-6.9 to 0.1) |
| 9 | 9 (0 – 17) | 17 (15 to 19) | -9.8 (-13 to -6.8) |
| 10 | 15 (0 – 40) | 17 (15 to 19) | -7.7 (-9.3 to -6.1) |
| 11 | 25 (11 – 40) | 20 (18 to 22) | -20 (-22 to -18) |
| 12 | 57 (19 – 84) | 12 (11 to 14) | -6.2 (-7.2 to -5.1) |
| 13 | 69 (31 – 91) | 15 (13 to 17) | -2.2 (-3.6 to -0.9) |
| 14 | 71 (39 – 91) | 15 (13 to 17) | -1.1 (-2.6 to 0.4) |

<sup>a</sup>For Germany and Netherlands only 2016 – 2019 were available
<sup>a</sup>Stringency in the non-pharmaceutical interventions at the population level (index 0 – 100) obtained from the Oxford COVID-19 Government Response Tracker

